# Efficacy and safety of novel probiotic formulation in adult Covid19 outpatients: a randomized, placebo-controlled clinical trial

**DOI:** 10.1101/2021.05.20.21256954

**Authors:** Pedro Gutiérrez-Castrellón, Tania Gandara-Martí, Ana Teresa Abreu, Cesar D. Nieto-Rufino, Eduardo López-Orduña, Irma Jiménez-Escobar, Carlos Jiménez-Gutiérrez, Gabriel López-Velazquez, Jordi Espadaler-Mazo

**Affiliations:** Centro de Investigación Translacional en Ciencias de la Salud, Hospital General Dr. Manuel Gea Gonzalez. Calz. De Tlalpan 4800, 14080 Ciudad de México, (CDMX), México; International Scientific Council for Probiotics, Tenango 22, 14340, Ciudad de México, (CDMX), México; Hospital Angeles Pedregal, Camino de Sta. Teresa 1055-S, 10700, Ciudad de México, (CDMX), México; DiagnoMol SA de CV. Camino Sta. Teresa No 13, 14110 Ciudad de México (CDMX), México; AB-Biotics SA (KANEKA Group), R&D Department, Av. De Torre Blanca 57, 08172 Sant Cugat, (Barcelona), Spain

## Abstract

**Background:** Probiotics have been proposed as adjuvants for Coronavirus Disease 2019 (Covid19) but randomized controlled trials (RCT) are lacking.

**Methods:** Single-center, quadruple-blinded RCT. Symptomatic Covid 19 outpatients (aged 18 to 60 years) with positive SARS-CoV2 nucleic acids test were randomized to active (n=150; ≥2×10^9^ colony-forming units (CFU) of probiotic strains *Lactiplantibacillus plantarum* KABP022, KABP023 and KAPB033, plus strain *Pediococcus acidilactici* KABP021) or placebo (n=150), take orally once daily for 30 days. Oral acetaminophen was allowed and controlled as co-intervention. Primary endpoint included: i) proportion of patients in complete remission (both symptoms and nucleic acids test) or progressing to moderate or severe disease with hospitalization; ii) death rate and duration on Intensive Care Unit (ICU). Safety was assessed in all patients. This study is registered at ClinicalTrials.gov (NCT04517422).

**Findings:** 300 subjects were randomized (median age 37.0 years [range 18 to 60], 161 [53.7%] women, 126 [42.0%] having known metabolic risk factors), and 293 completed the study (97.7%). Remission was achieved by 78 of 147 (53.1%) in the active group compared to 41 of 146 (28.1%) in placebo (P<0.0001; ARR=25.0% [95%CI 14.1-35.9%]), still significant after multiplicity correction for the primary endpoint. No hospitalizations or deaths occurred during the study, precluding the assessment of efficacy on these endpoints. No serious adverse events occurred during the study. Replication studies with this probiotic formula are warranted.

## INTRODUCTION

Severe acute respiratory syndrome coronavirus 2 (SARS-CoV2), first identified in December 2019 in Wuhan, China, is the causative agent of Coronavirus Disease 2019 (Covid19) global pandemic.^1^ SARS-CoV2 infection can range from asymptomatic to Acute Respiratory Distress Syndrome (ARDS) and death. Most symptomatic patients typically display mild to moderate symptoms, even despite significant viral loads, ^2^ and their condition can be managed on an outpatient basis. Typical symptoms include dry cough, fever, shortness of breath, body aches, headache, fatigue, diarrhea, nausea, anosmia and ageusia. ^3^ Several risk factors have been identified for severe Covid19, including diabetes, hypertension, older age, being male and ethnicity. ^4^ However, no therapies have been approved for outpatients with Covid19 disease to date.

Recent evidence suggests the existence of a crosstalk between the gastro-intestinal tract and respiratory system, along with their respective microbiomes, referred to as the gut-lung axis (GLA). Intestinal bacteria can potentially achieve distal effects on lung homeostasis through training of immune cells in the gut epithelium and their subsequent migration to the pulmonary epithelium, and/or through the permeation into the bloodstream of bacterial metabolites having distal immunomodulatory effects. ^5,6^

Probiotics are defined as “live microorganisms that when administered in adequate amounts, confer a health benefit on the host”, and this definition entails the requirement of well-conducted studies in humans in the specified health indication. ^7^ Past meta-analyses have suggested probiotics may have a role in respiratory infections such as cold and influenza, but significant heterogeneity was noted between individual trials. ^8,9^ This is to be expected, since many probiotic effects are strain-specific, particularly immune effects. ^7,10–12^ Besides, safety issues are often poorly reported in probiotic trials, ^13^ further hampering a proper assessment of harms vs. benefits. Nevertheless, probiotics have been proposed for Covid19^14^ although no randomized, placebo-controlled trials have been published to date.

Most current probiotics are lactic acid bacteria. Such bacteria are often associated with the dairy industry, but in fact several species are present in the phyllosphere and endosphere of many plants (*i*.*e*. living on the surface or inside plants). Transiting through the gut of herbivores and omnivores is part of the lifecycle of such nomadic bacteria. ^15^ *Lactiplantibacillus plantarum* (formerly known as *Lactobacillus plantarum*) and *Pediococcus acidilactici* are two such species of lactic acid bacteria.

In this study we evaluated the efficacy and safety of a probiotic composed of three *L. plantarum* stains (KABP022, KABP023 and KABP033) and one *P. acidilacti strain* (KABP021) in symptomatic Covid19 outpatients. To that end, we used both laboratory and patient-reported outcomes. Compared to placebo, we show that a 30-day intervention with this probiotic resulted in increased Covid19 remission at study endpoint (requiring both symptom clearance and negative nucleic acids test). Shortening of the duration of several symptoms, changes in humoral immunity against SARS-CoV2 (spike-binding IgM and IgG antibodies) and changes in lung abnormalities (as per chest X-ray imaging) were also achieved.

## METHODS

### Study Design

Randomized, quadruple-blinded (patient, caregiver, investigator and outcomes assessors), randomized, placebo-controlled, parallel-arms clinical trial, in outpatient subjects with recently diagnosed Covid19. The trial complied with the Declaration of Helsinki and applicable local regulations and was approved by the Research Ethics Committee of Hospital General Dr. Manuel Gea Conzalez of Mexico City (approval number 12-120-2020, ClinicalTrials.gov identifier: NCT04517422). All participants provided written informed consent before entering the study. (CONSORT2010 checklist in Supplementary Material).

### Participants

Outpatient subjects, 18 to 60 years old, with diagnosis of SARS-CoV2 infection confirmed by Reverse-Transcriptase Quantitative Polymerase Chain Reaction (RT-qPCR) (Supplementary Methods), and positive for Covid19 symptoms were recruited at Hospital General Dr. Manuel Gea Gonzalez, a tertiary referral hospital in Mexico City (Mexico). Five Covid19 symptoms were considered for enrolment: cough, headache, fever (>37.5°C), muscular pain and shortness of breath (at least one was required). Other inclusion criteria were symptom onset ≤7 days prior to recruitment and peripheral oxygen saturation (SpO_2_) ≥ 90%. Investigators reviewed symptoms, risk factors, and other non-invasive inclusion and exclusion criteria prior to enrollment (full list of inclusion/exclusion criteria, as well as reason for study drop-out appears in Supplementary Methods).

### Randomization and Masking

Subjects were randomized in a one-to-one proportion in blocks of size six without stratification, using a randomization list generated with the Sealed Envelope web service (https://www.sealedenvelope.com/), by a study site pharmacist not participating in the study. Participants were enrolled and assigned to study groups by their caregivers at the study site. Study products (active or placebo) were given in coded, anonymous boxes, and subjects, caregivers, investigators and outcome assessors were unaware of the treatment allocation. All personnel involved in the study remained unaware of subjects allocation until randomization list was opened in the presence of a witness on Feb 3^rd^, 2021, once all study subjects had completed the intervention and primary statistical analysis had been performed.

### Study Products

The active product consisted of a blend of four strains of freeze-dried lactic acid bacteria: *Lactiplantibacillus plantarum* KABP033 (CECT30292), *L. plantarum* KABP022 (CECT7484), *L. plantarum* KABP023 (CECT7485)) and *Pediococcus acidilactici* KABP021 (CECT7483), in a ratio of 3:1:1:1 colony-forming units (CFU), respectively, with a maltodextrin carrier. This blend was prepared in HPMC (hydroxymethyl propyl cellulose) hard shell capsules at >2×10^9^ total CFU/capsule. Placebo product consisted of HPMC capsules containing the maltodextrin carrier only. Active and placebo samples were supplied by Kaneka AB-Biotics SA (Barcelona, Spain) and were indistinguishable in form, color, and taste.

Identity of the four strains in the active product batch and microbial quality of active and placebo batches were verified, and the strains genomes were confirmed to be devoid of antimicrobial resistance genes (Supplementary Methods). Active sample was monitored for conformance to specification (≥2×10^9^ CFU/capsule) throughout the study in stability chamber (25 ± 2°C, 60 ± 5% relative humidity) by ISO17025-accredited company Silliker Iberica (Barcelona, Spain, part of Merieux Nutrisciences Group).

### Procedures

The study was scheduled across 3 study site visits: day 0 (visit 1, screening), day 15 ± 1 (visit 2) and day 30 ± 1 (visit 3, end of intervention). Demographic and clinical data were collected on day 0. Study subjects were given the study product on visit 1 and were instructed to store it at room temperature (max. 25°C) and take one treatment capsule (active or placebo) daily orally, from day 1 to day 30, 20 minutes before breakfast. Study subjects were also given access to a web-based electronic diary for daily recording of symptoms, body temperature and peripheral oxygen saturation. With the aim of minimizing heterogeneity in reported data, a YBK303 pulsoxymeter (Yobekan Medical Equipment, Henan, China) and a Harbin infrared thermometer (Harbin Xiande Technology Development Co, Harbin, China) were provided to each subject for at home use during the study.

On study visits, subjects were assessed for severity of Covid19 using WHO Clinical Progression Scale for Covid19,^16^ and received chest pulmonary X-ray. Venous blood and nasopharyngeal samples were also taken for serum biomarker and SARS-CoV2 RT-qPCR analysis, respectively (Supplementary Methods). Fecal samples for microbiome 16S rRNA analysis were collected on visits 1 and 3 with GUT-OMR200 kit (DNAgenotek, Ottawa, Canada). Study subjects were also contacted by phone on days 5, 10, 20 and 25 (all ± 1 day) by a physician as part of outpatient follow-up. Calls were also used to encourage adherence to study procedures.

On demand acetaminophen (500 mg/dose, up to three times a day) was allowed as concomitant medication for Covid19 symptoms, and its use was reported in patient diary.

### Outcomes

The primary efficacy endpoint was the frequency of subjects who progressed from ambulatory mild disease to either remission, hospitalized moderate disease; or hospitalized severe disease, at the end of the 30 days intervention, according to WHO Clinical Progression Scale.^16^ Particularly, remission was defined as a negative RT-qPCR (viral clearance) plus complete resolution of all the five symptoms considered at study entry (fever, cough, headache, body aches and shortness of breath). Mortality rate and length of stay in Intensive Care Unit (ICU) at the end of the 30 days intervention were also considered co-primary endpoints, thus totaling five co-primary endpoints.

The prespecified secondary efficacy endpoints included: i) SARS-CoV2 viral load evaluated by RT-qPCR; ii) Plasma SARS-CoV2 spike protein-specific IgG and IgM levels; iii) duration of each of five core Covid19 symptoms from the start of the intervention in patient diaries: fever (>37.5°C), cough, headache, body aches and shortness of breath; iv) high sensitivity C-reactive protein (hsCRP) and D-Dimer; and v) lung abnormalities measured by chest X-ray, classified according to Brixia score.^17^ Additional prespecified secondary endpoints included: i) gastrointestinal symptoms; and ii) fecal microbiome evaluated by 16S rRNA analysis. At the time of writing, evaluation of these two additional endpoints is still ongoing and will be reported elsewhere.

Originally, remission was not considered for the primary endpoint. However, as the study progressed, it became apparent that no hospitalizations were occurring, and thus chances of observing a sizeable number of hospitalizations at the end of the study was low. Therefore, the principal investigator proposed adding remission (defined as negative RT-qPCR plus symptomatic remission) to primary outcomes to the Research Ethics Committee, while keeping other predefined categories to minimize changes. The principal investigator also proposed including the duration of specific Covid19 symptoms, namely fever (>37.5C), cough, headache, shortness of breath and body aches) as prespecified secondary endpoints, since these symptoms were already being recorded in the patient’s diary and were used by doctors to track the overall symptomatic evolution of patients. These changes were approved by the Research Ethics Committee on Jan 27^th^, 2021 (before study unblinding).

Finally, exploratory, non-prespecified (post-hoc) endpoints included: i) significance of the primary endpoint when splitting the population according to age (less than 50 years old vs 50 years and older), sex (male vs female), metabolic comorbidity (diabetes, hypertension or obesity vs none), viral load at baseline (below vs above median value at baseline) and time from symptom onset (one to four days vs five or more days); ii) time to symptomatic resolution, defined as the disappearance of all five core Covid19 symptoms according to patient diaries (fever, headache, cough, body aches and shortness of breath); and iii) number of days of use of acetaminophen, of loss of taste (ageusia) and of loss of smell (anosmia).

A treatment-emergent adverse event (AE) was defined as any event that first occurred or worsened in severity after the initiation of the intervention, akin to other trials in Covid19 outpatients. ^18^ A serious AE (SAE) was defined when causing hospitalization, persistent disability or incapacitation, or death. Reporting of adverse events was monitored by phone calls (days 5, 10, 20 and 25), as well as the study site visits (days 0, 15 and 30). Finally, treatment-emergent increases in serum hsCRP reaching >3 mg/L were also considered in safety analysis.

### Sample Size

No published data could be found regarding the risk of Covid19 progression from mild to moderate or severe disease in Mexico, but early estimates based on local experience ranged 27 to 67%. Taking the average value (47%) and aiming at detecting a relative reduction of at least 35% with a two-sided alpha = 5% and power = 80% resulted in 150 subjects per study arm after rounding up.

### Statistical Analysis

Analyses were performed according to assigned randomization group, without any data imputation for missing values. Significance testing for the primary endpoint was performed by Pearson’s Chi-squared test, while binomial logistic regression was used when adjusting for baseline covariates in sensitivity analysis. Pearson’s Chi-squared test was also used to assess differences between groups in other binary variables, while differences in continuous variables at baseline and in days of symptoms were assessed with Mann-Whitney test. Differences between groups across days 0, 15 and 30 in SARS-CoV2 viral load, SARS-CoV2-specific IgM and IgG, hsCRP, D-Dimer and Chest-X ray Brixia score were assessed by mixed-effects models for repeated measures (MMRMs), with study group and visit as fixed factors, a group-by-visit interaction term and unstructured covariance matrix. Finally, time to overall symptom resolution was assessed by Kaplan-Meyer survival analysis. All statistical analyses were performed with the SPSS program v.24 (IBS Corp., Armonk, US). Differences were considered significant at two-sided *P* < 0.05. For primary endpoint, a Bonferroni-type correction for multiplicity was applied, accounting for the five prespecified comparisons, resulting in a more stringent significance threshold of P < 0.01.

## RESULTS

### Participants

Of the 300 patients randomized, 293 completed the study between August 26^th^ and December 10^th^ 2020 and were available for primary analysis, while 7 were lost to follow-up (3 in active and 4 in placebo, CONSORT Flowchart in Figure 1). Median patient age was 37.0 years (range 18 to 60), 161 (53.7%) were women and 126 (42.0%) had known metabolic risk factors (BMI ≥ 30, diabetes and/or hypertension). Patients in active and placebo groups were well balanced at baseline (Table 1), except for the following variables: active group had a higher incidence of lung abnormalities and of type II obesity, and lower SpO2 at baseline, while placebo group had higher incidence of type I obesity and of shortness of breath. Said variables were thus considered for sensitivity analysis of the primary endpoint.

**Table 1.** Demographic data of the subjects at baseline.

|  | Active (n=150) | Placebo (n=150) |
| --- | --- | --- |
| Age (years) [median, range] | 34 [19-60] | 39 [18-59] |
| Sex (female) [n, %] | 82 [54.7%] | 79 [52.7%] |
| Body Mass Index (BMI, kg/m<sup>2</sup>) [median, range] | 27.5 [20.1-39.3] | 29.4 [22.0-39.5] |
| • Of which class I obesity (BMI 30 to <35) [n, %] | 31 [20.7%] | 72 [48.0%] |
| • Of which class II obesity (BMI 35 to <40) [n, %] | 16 [10.7%] | 0 [0.0%] |
| Smoker (yes) [n, %] | 22 [14.7%] | 20 [13.3%] |
| Diabetes (yes) [n, %] | 15 [10.0%] | 16 [10.7%] |
| Arterial hypertension (yes) [n, %] | 28 [18.7%] | 31 [20.7%] |
| Poly-medicated (yes) [n, %] | 24 [16.0%] | 18 [12.0%] |
| Use of acetaminophen (yes) [n, %] | 83 [55.3%] | 70 [46.7%] |
| Days from symptom onset [median, range] | 4 [1-7] | 4 [1-7] |
| Fever (yes) [n, %] | 100 [66.7%] | 115 [76.7%] |
| Cough (yes) [n, %] | 138 [92.0%] | 133 [88.7%] |
| Headache (yes) [n, %] | 134 [89.3%] | 127 [84.7%] |
| Shortness of breath (yes) [n, %] | 42 [28.0%] | 64 [42.7%] |
| Body aches (yes) [n, %] | 94 [62.7%] | 97 [64.7%] |
| Loss of taste (yes) [n, %] | 55 [36.7%] | 62 [41.3%] |
| Loss of smell (yes) [n, %] | 57 [38.0%] | 66 [44.0%] |
| Diarrhea or loose stools (yes) [n, %] | 55 [36.7%] | 69 [46.0%] |
| Nausea (yes) [n, %] | 46 [30.7%] | 47 [31.3%] |
| Abdominal pain (yes) [n, %] | 22 [14.7%] | 16 [10.7%] |
| Lung abnormalities (yes) [n, %] | 72 [48.0%] | 48 [32.0%] |
| Peripheral oxygen saturation (SpO<sub>2</sub>, %) [median, range] | 90 [90-95] | 91 [90-93] |
| SARS-CoV2 nasopharyngeal viral load (log10 of copies/mL) [median, range] | 6.8 [6.0-7.0] | 6.8 [6.1-7.5] |
| SARS-CoV2 spike protein-specific IgM (seropositive) [n, %]* | 150 [100%] | 150 [100%] |
| SARS-CoV2 spike protein-specific IgG (seropositive)<br>[n, %]* | 36 [24.0%] | 31 [20.7%] |
| hsCRP (mg/L) [median, range] | 3.24 [0.93-5.43] | 3.43 [1.21-6.98] |
| D-Dimer (mg/L) [median, range] | 2.02 [0.29-3.45] | 2.04 [0.46-3.82] |

\*) Defined as per manufacturer instructions.

**Figure 1.**
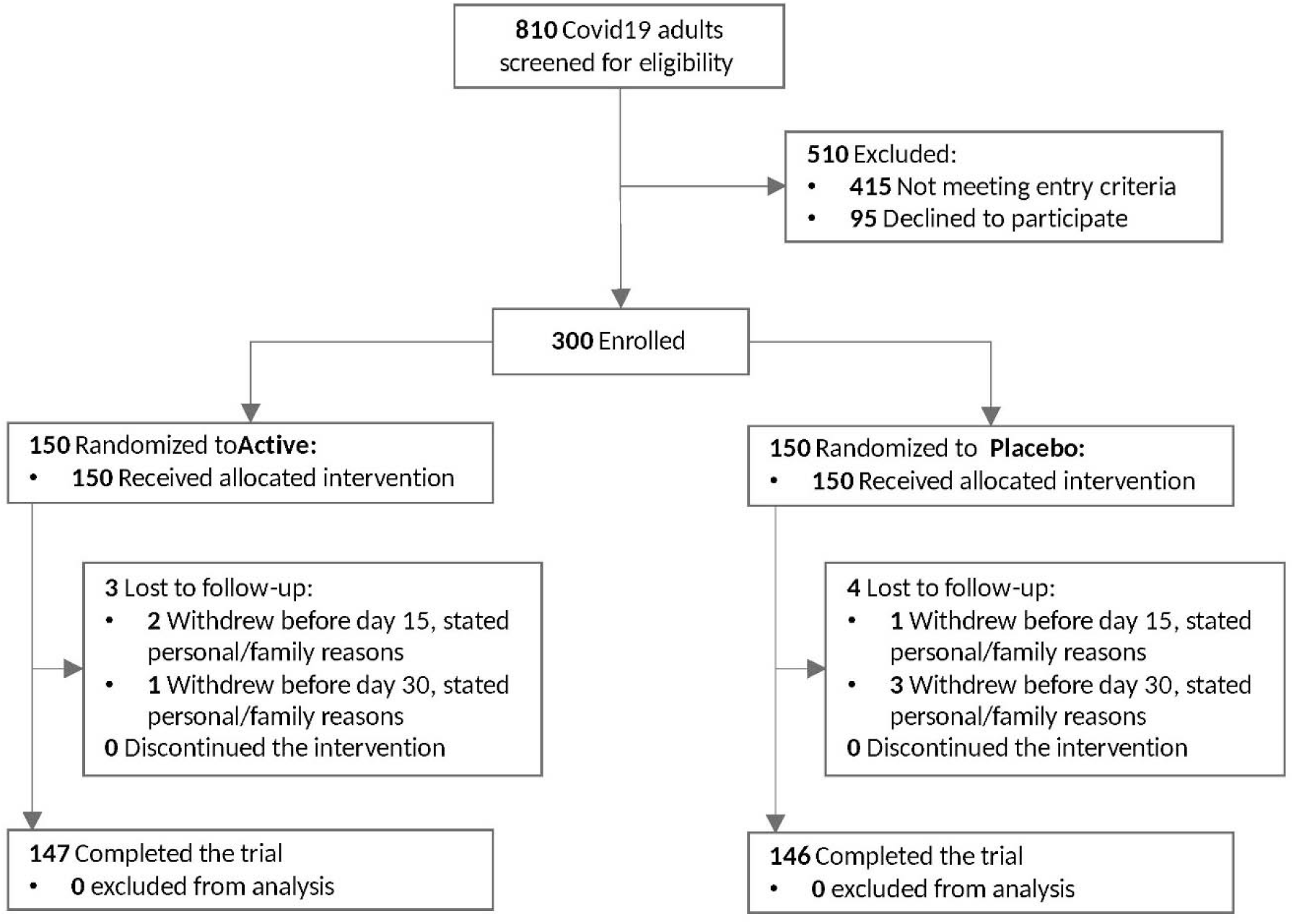
Patient enrollment and treatment assignment to probiotic (≥2×10^9^ CFU, active arm) or control (placebo arm) among symptomatic Covid19 outpatients (CONSORT 2010 Flowchart).

**Figure 2.**
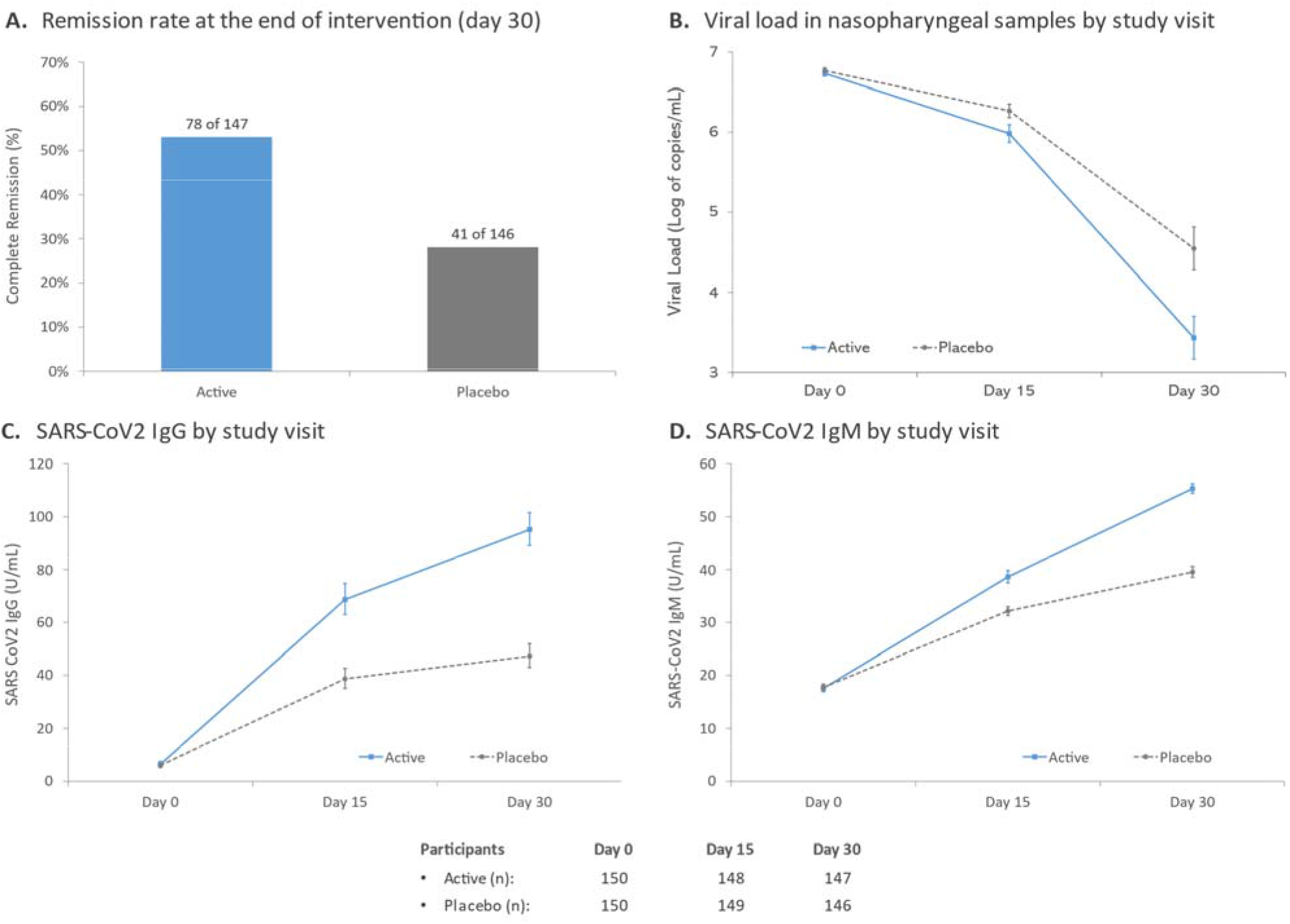
A) Remission rate (symptom clearance for at least 24h plus negative RT-qPCR) on day 30. B) Evolution of mean viral load (as base 10 logarithm of viral copies/mL), as measured by RT-qPCR. C) Evolution of geometric mean serum levels of SARS-CoV2 spike protein-specific IgM. D) Evolution of geometric mean serum levels of SARS-CoV2 spike protein-specific IgG. Error bars denote 95%CI of the geometric means.

### Primary Outcomes

Remission rate on day 30 was 53.1% (78 of 147) in the active group compared to 28.1% (41 of 146) in the placebo group (P < 0.0001; ARR: 25.0% [95%CI 14.1-35.9%]; OR: 2.90 [95%CI 1.78-4.70]) (Figure 1A and Table S1). This p-value remained significant at the multiplicity-corrected threshold of P = 0.01. No hospitalizations, ICU admissions or deaths occurred during the study, preventing the assessment of remaining primary endpoints. Thus, no differences could be found regarding the rate of hospitalization (moderate or severe disease) or death, nor the duration of ICU stays.

### Secondary Outcomes

Among secondary endpoints, patients in active group reported less days of fever, cough, headache, body aches and shortness of breath, the effect in symptoms other than fever being dependent on baseline status (Table 2). Patient diary compliance was high, with only 17 of 300 subjects (11 in active and 6 in placebo, P = 0.213) failing to report 100% complete diaries. Repeated measures analysis indicated significantly larger reduction of nasopharyngeal viral load in active vs. placebo (P < 0.0001; Figure 1B), as well as higher increase of SARS-CoV2-specific IgG and IgM in serum (both P < 0.0001; Figure 1C and D). These differences on viral load, IgM and IgG were significant both on day 15 and day 30. Active treatment also reduced serum levels of both hsCRP and D-Dimer compared to placebo, this effect being pronounced on day 15 (both P < 0.0001) but waning out on day 30 (Supplementary Figures S1A and S1B). Finally, among those subjects with lung abnormalities, active treatment resulted in a significantly better improvement of the radiographic scoring both on days 15 and 30 (both P < 0.0001; Supplementary Figure S2).

**Table 2.** Symptom duration, split by baseline status for each symptom (yes/no), indicated as median days [range]. Number of subjects in each subgroup are indicated within parentheses below.

|  | Baseline status | Active | Placebo | P-value |
| --- | --- | --- | --- | --- |
| Fever | Yes | 2 [0-11]<br>(n=100) | 5 [3-17]<br>(n=115) | < 0.0001 |
|  | No | 2 [0-13]<br>(n=50) | 4 [3-14]<br>(n=35) | < 0.0001 |
| Cough | Yes | 10.5 [0-15]<br>(n=138) | 14 [0-26]<br>(n=133) | < 0.0001 |
|  | No | 0 [0-8]<br>(n=12) | 0 [0-10]<br>(n=17) | 0.2377 |
| Headache | Yes | 7 [0-13]<br>(n=134) | 12 [0-26]<br>(n=127) | < 0.0001 |
|  | No | 0 [0-0]<br>(n=16) | 0 [0-1]<br>(n=23) | 0.4043 |
| Shortness of<br>breath | Yes | 2.5 [0-9]<br>(n=42) | 5 [0-13]<br>(n=64) | 0.0002 |
|  | No | 0 [0-0]<br>(n=108) | 0 [0-0]<br>(n=86) | 1.0000 |
| Body aches | Yes | 3 [0-9]<br>(n=94) | 7 [0-14]<br>(n=97) | 0.0001 |
|  | No | 0 [0-6]<br>(n=56) | 0 [0-7]<br>(n=53) | 0.5937 |

### Exploratory Outcomes (post-hoc)

Sensitivity analyses were performed to assess the robustness of this result, since imbalances were observed for some variables at baseline. Upon adjusting for said imbalances, active group remained significantly associated to remission (P < 0.0001; OR: 2.98 [95%CI: 1.77-5.03], Table S2). Significance for the primary endpoint was also retained in all the trial subpopulations assessed (split by age, sex, presence of metabolic comorbidity, baseline viral load and days from symptom onset; Table S3). Median time to overall symptom resolution was significantly shorter in active than placebo group (Supplementary Figure S3). Finally, median number of days of loss of taste, of loss of smell and of use of acetaminophen were also lower in active group than placebo (Supplementary Table S4).

### Safety

Serious adverse events (SAEs) occurred in 0% (0 of 300) of study subjects (Table 3). Treatment-emergent adverse events (AEs) were reported in 41 (27.3%) and 63 (42.0%) subjects of active and placebo groups, respectively, the most frequent being emergent fever, cough, body aches, pain when swallowing and conjunctivitis. Incidence of body aches was higher in active than placebo group, but the difference did not reach statistical significance (10 of 150 vs 5 of 150; p = 0.186). Incidence of other AEs was generally higher in placebo than active group, and thus most AEs could be likely due to natural symptoms flares in Covid19 disease. Besides, hsCRP treatment-emergent increases in hsCRP levels reaching values above 3 mg/L were found in 2 subjects in placebo group and none in active group. No subjects in the study displayed hsCRP levels above 10 mg/L.

**Table 3.** Treatment-emergent adverse events (AEs) observed during the 30-day study period. A treatment-emergent adverse event was defined as an event that first occurred or worsened in severity after study initiation, or isolated events that occurred 14 or more days after initial disappearance of symptoms and were followed by a negative RT-qPCR. AEs were reported by the participant in their patient diary, or, when appropriate, by the participant’s legally authorized representative. A serious AE (SAE) was defined when causing hospitalization, persistent disability or incapacitation, or death.

|  | Active<br>(*n* = 150) | Placebo<br>(*n* = 150) |
| --- | --- | --- |
| Patients with $\geq 1$ AE | 41 (27.3%) | 63 (42.0%) |
| Patients with $\geq 1$ SAE | 0 | 0 |
| AEs occurring anytime during the study |  |  |
| • Fever ($\geq 37.5^{\circ}\text{C}$) | 7 (4.6%) | 19 (12.73%) |
| • Poor appetite | 12 (8.0%) | 11 (7.3%) |
| • Cough | 5 (3.3%) | 12 (8.0%) |
| • Body aches | 10 (6.7%) | 5 (3.3%) |
| • Pain when swallowing | 4 (2.7%) | 10 (6.7%) |
| • Conjunctivitis | 4 (2.7%) | 8 (5.3%) |
| • Shortness of breath | 0 | 9 (6.0%) |
| • Headache | 2 (1.3%) | 7 (4.7%) |
| • Diarrhea | 4 (2.7%) | 3 (2.0%) |
| • Chest oppression | 0 | 3 (2.0%) |
| • Hard stools | 0 | 2 (1.3%) |
| • Nausea | 0 | 1 (0.7%) |
| • Feeling of bowel movement urgency | 0 | 1 (0.7%) |

## DISCUSSION

In this blinded, randomized study in Covid19 outpatients, the probiotic formula achieved a significant effect on improving remission rate against placebo (Number Needed to Treat of 4). No patients were hospitalized or died during the intervention, preventing assessment of other co-primary endpoints (frequency of progression to hospitalization, mortality ration, duration of ICU stay). In this regard, our entry criteria preventing the recruitment of subjects older than 60 years or having SpO2 below 90% likely resulted in a population with markedly less hospitalization risk than initially expected. In fact, recent RCT in similar Covid19 outpatients, without the age and SpO2 limits used in our study, found an incidence of hospitalization or emergency department visit of just 6% in the placebo group. ^18,19^ Nevertheless, the significance achieved in remission (p < 0.0001) was well below the Bonferroni-corrected threshold for multiplicity of co-primary endpoints and was also robust to multivariate adjustment for baseline confounders. Moreover, post-hoc analyses showed the effect was consistent across study subpopulations based on age, sex, metabolic comorbidities, viral load at baseline and days from symptom initiation (all with p < 0.05). Importantly, the outcome of remission required both the disappearance of all five prespecified symptoms (fever, cough, headache, body aches and shortness of breath) and a negative RT-qPCR result (*i.e*. less than one viral copy per reaction volume).

Among the secondary endpoints, the probiotic intervention was observed to significantly shorten the duration of each of the five prespecified, non-intestinal Covid19 symptoms, the effect being dependent on baseline status for most symptoms. Probiotic intervention was also found to lower viral load, lower the scoring of lung abnormalities and increase SARS-CoV2 IgM and IgG in a significant manner. Most symptomatic Covid19 patients typically display mild to moderate symptoms, and their condition can be managed on an outpatient basis. However, few RCTs have found therapies effective at increasing viral clearance and/or shortening symptom duration in Covid19 outpatients so far^18–24^. To our best knowledge, this is the first RCT to report such an effect for a probiotic in Covid19, since a previous probiotic trial (with different strains) was open-label and non-randomized. ^25^

Within two weeks of symptom onset, virus-specific antibodies against SARS-CoV2 start to increase, followed by decay of IgM and IgA while IgG can remain high for weeks and months. ^26^ In our study, spike protein-specific IgM were higher than IgG at baseline but this trend was reversed already on day 15. Probiotic intake resulted in further increase in SARS-CoV2-specific IgM and especially a twofold increase in IgG compared to placebo, suggesting a crosstalk between this probiotic and adaptive humoral immunity to a respiratory pathogen. The effect on multiple objective endpoints such as virus-specific humoral immunity, lung abnormalities and viral load against placebo supports a true effect of this probiotic on the GLA. Of note, this effect could be independent of significant changes in microbiome composition and due solely to a crosstalk between the probiotic and the host immune system. Microbiota-derived molecules, such as short chain fatty acids, tryptophan metabolites, toll-like receptor (TLR), nucleotide-binding oligomerization domain (NOD) ligands, can act directly on local intestinal tissue cells, but also penetrate into circulation to tune immune cells in peripheral tissues. ^5^ Besides, some bacterial surface proteins are directly recognized by different types of antigen-presenting cells, ^11,12^ which could result in systemic effects via migration of primed T-cells. In this regard, the genomes of *L. plantarum* strains KABP022, 023 and especially KABP033 code for one such protein (Supplementary Table S5). In Covid19, additional mechanism in GLA cross-talk could involve direct modulation of angiotensin converting enzyme (ACE) 2 expression in the gut: Intestinal ACE2 is known to be affected by SARS-CoV2 infection, lack of ACE2 is known to reduce tryptophan absorption via the neutral amino acid transporter B^0^AT1, ^27^ and tryptophan-derived metabolites play important roles in gut homeostasis and systemic immunity. ^28^ Future studies should aim at unravelling the precise mechanisms underlying the effect of the strains used in this study on adaptive immunity and the GLA.

Treatment-emergent adverse events were characterized as in recent Covid19 trials^18^ and the results of this study highlight the safety of this probiotic formulation in Covid19 outpatients. Of note, despite the stimulating effect on SARS-Cov2-specific immunoglobulins, general inflammation status did not seem to increase, according to hsCRP measurements. Human supplementation with probiotic microorganisms is generally considered to be safe, based on the history of use of probiotics in foods, and is recognized as such for most probiotic strains by regulatory authorities. ^13,29^ Conversely, their use in patients with severe disease remains controversial due to concerns of bacteremia by lactic acid bacteria or microbial contaminants, especially immunosuppressed patients or those in intensive care units (ICU), ^13^ although known cases of probiotic-associated bacteremia are mostly associated to strains not used in this study. ^13,30,31^ Moreover, transmission of antibiotic resistance genes from probiotics to pathogenic bacteria has not been demonstrated in vivo, but remains a valid concern. In this regard, the genomes of the four strains used in this study were confirmed to be devoid of transmissible antibiotic resistance genes, and potential microbial contaminants were analyzed in study product. However, additional studies should be conducted before the use of this probiotic can be recommended to hospitalized patients with severe Covid19.

This study has several limitations worth mentioning. First, no patients older than 60 years old were included in the study. However, the consistency of the effect across age subpopulations when splitting the study sample at the 50 years-old cutoff suggests the effects of this probiotic are not limited to young adults, but additional studies in older populations are warranted. Second, all subjects in the study were of Hispanic ethnicity, which has been associated to higher mortality in Covid19. ^32^ In our study, reduction of viral load in placebo group was slower than reported in similar studies where Hispanic subjects accounted for 50% or less of the study population. ^18,19^ Accordingly, our study population could be regarded as more challenging, but replication studies should be performed in more diverse populations. Third, the 16S stool microbiome analysis is still ongoing at the time of writing. Microbiome analysis could provide clues about the mechanism of action of the probiotic formula used in this trial and/or reveal if baseline enterotype influences the effectiveness of the probiotic. However, microbiome changes are not required for a probiotic to be effective since activity could be due to crosstalk between the probiotic strains per se and the host’s immune system. Results of microbiome analysis and digestive symptoms will be jointly reported in future publications. Fourth, the SARS-CoV2 strains affecting patients in this study were not determined.

In our view, this study also has some strengths worth commenting. First, concomitant on-demand use of acetaminophen was allowed in this study (as a common practice for Covid19 outpatients) and recorded daily in patients’ diaries, thus ruling out a confounder effect. Second, strain composition of the formula was verified by PCR, and the method is provided to facilitate replication in future studies. Third, stability of the probiotic composition was verified at the end of the study by an independent analysis lab. Fourth, phone calls were scheduled across the study to check on patients and remind of study procedures, resulting in low drop-out (7 of 300, *i*.*e*. 2.33%) and high patient diary compliance. Fifth, quadruple blinding and use of objective measures support the reliability of the results.

Future replication studies should take into account two considerations. On the one hand, any replication study must ensure the very same strains are used and at the same dosing, not other *L. plantarum* and *P. acidilactici* strains. To that end, we provide a method for the identification of the strains used herein. On the other hand, the putative mechanisms of action of probiotics on immune homeostasis probably require a build-up time. In our view, probiotics should not be expected to act as fast as antipyretics or corticosteroids, and clinical trials should be designed accordingly.

In conclusion, among non-hospitalized patients with mild to moderate COVID-19 illness, treatment with a probiotic composed of strains *P. acidilactici* KAP021 and *L. plantarum* KABP022, KABP023 and KABP033 at a total dose of ≥ 2 × 10^9^ CFU/day was associated with a statistically significant increase in remission on day 30 compared to placebo. Effect on hospitalization, duration of ICU stay and mortality could not be assessed because of lack of occurrences during the study. Significant effects on symptom duration, viral load, lung abnormalities and SARS-CoV2-specific IgM and IgG were also observed, and the probiotic formula was well tolerated. In or view, these results warrant replication studies with this probiotic formula.

## CONTRIBUTIONS

PGC, JEM and ATAA conceptualized and designed the study. PGC was the study coordinator. TGM, CDNR and IJE performed patient procedures, and ELO and GLV performed laboratory analyses. CJG collected the data and curated the data together with PGC. PGC and JEM performed formal statistical analyses and wrote the manuscript. ATAA critically revised the manuscript.

## Supporting information

Appendix (supplementary materials, supplementary results and CONSORT checklist)

ICMJE Conflict of Interest Statements

## Data Availability

Deidentified individual patient data (IPD), together with data dictionary defining each field in the set, will be made public upon any formal requests with a defined analysis plan. To make a request, please contact the corresponding author. Decision to make IPD publicly available was taken after initial trial registration.

## ACKNOWLEDGMENTS

Our sincere thanks to thank Paola Juarez-Valdez, Pamela Paez-Melendez, Yarumi Espinosa-Hernandez and Nidia Gómez-Ramirez for the coordination and nursing activities on the project. This study was funded by AB-Biotics S.A (Barcelona, Spain), a member of the KANEKA Group (Japan).

