## Appendix (supplementary materials, supplementary results and CONSORT checklist) for "Efficacy and safety of novel probiotic formulation in adult Covid19 outpatients: a randomized, placebo-controlled clinical trial"

### Supplementary Methods

##### Complete list of Inclusion and Exclusion criteria, and Reasons for Study Withdrawal

| | |
|---|---|
| Inclusion | • Age between 18 and 60 years old, both included.<br>• Positive RT-qPCR for the SARS-CoV2 RdRP gene (*i.e.* ≥1 copy per PCR reaction volume).<br>• At least one of the following symptoms, with symptom onset within ≤7 days of study entry: fever (*i.e.* >37.5ºC), cough, headache, body aches and shortness of breath.<br>• Peripheral oxygen saturation ($SpO_2$) ≥90%.<br>• Ability to understand study procedures.<br>• Providing informed consent. |
| Exclusion | • Uncontrolled diabetes (*i.e.* HbA1c > 8%).<br>• Uncontrolled systolic hypertension (*i.e.* >160 mmHg).<br>• Body Mass Index (BMI) ≥40 $Kg/m^2$.<br>• Known coagulopathy.<br>• Acute pancreatitis.<br>• Chronic diarrhea, chronic constipation or Inflammatory Bowel Disease (IBD).<br>• Currently immunosuppressed due to cancer treatment, transplant, HIV infection or treatment of autoimmune disease.<br>• Severe respiratory disease (asthma, chronic obstructive pulmonary disease, cystic fibrosis).<br>• Ongoing severe seasonal allergies.<br>• Known glucose 6-phospate dehydrogenase deficiency.<br>• Probiotic or antibiotic use within 2 days before entering the study.<br>• Pregnancy or lactation. |
| Reasons for Study Withdrawal | • Severe non-compliance to Study Protocol procedures, as judged by the Investigator<br>• Subject withdraws informed consent or is lost to follow-up<br>• Significant adverse effect (AE) posing a risk for the subject, as judged by the Investigator.<br>• Onset of a comorbidity described as exclusion criteria during the study |

**Whole-Genome Search of Antimicrobial Resistance Genes**

Complete genomes of strains *P. acidilactici* KABP021 (CECT7483), *L. plantarum* KABP022 (CECT7484) and *L. plantarum* KABP023 (CECT7485) had been previously obtained by MiSeq sequencing (Illumina Inc., San Diego, US) and assembled with MegaHit software (https://github.com/voutcn/megahit) while complete genome of novel strain *L. plantarum* KABP033 (CECT30292) was obtained by PacBio Sequel II sequencing (Pacific Biosciences Inc. Menlo Park, US) and assembled to obtain supercontigs (long read sequence data) with Flye software (https://github.com/fenderglass/Flye). The obtained genomes were double-checked against the most up-to-date version of two dedicated databases, using default settings:

- The Comprehensive Antibiotic Resistance Database (CARD, https://card.mcmaster.ca/), a bioinformatic database of resistance genes, their products, and associated phenotype.[1]
- The ResFinder database (https://cge.cbs.dtu.dk/services/ResFinder/) to identify putative acquired antimicrobial resistance genes and/or chromosomal mutations.[2]

Putative antimicrobial resistance genes were found in none of the four strains genomes:

| Strain | Genome Size (bp) | Contigs | GC (%) | Hits in CARD | Hits in ResFinder |
|---|---|---|---|---|---|
| Pa KABP021 | 1,973,667 | 24 | 42.1 | None | None |
| Lp KABP022 | 3,368,099 | 44 | 44.4 | None | None |
| Lp KABP023 | 3,216,006 | 41 | 44.5 | None | None |
| Lp KABP033 | 3,322,082 | 4 | 44.4 | None | None |

**Verification of Strain Identity in Active Product**

The content of 1 capsule was resuspended in 1 mL of phosphate-buffered saline (PBS). DNA from PBS-resuspended samples was extracted using QIAamp DNA Mini Kit (Qiagen, Hilden, Germany), following manufacturer's instructions. PCR experiments were performed using strain specific primers (see table below) and MyTaq™ DNA Polymerase (BioLine, Meridian Bioscience, US) with the following parameters: initial denaturation step (1 min. at 95ºC); 35 cycles comprising denaturation (15 sec. at 95ºC), annealing (15 sec. at 60ºC) and elongation (10 sec. at 72ºC); and a final elongation step (7 min. at 72ºC). The composition of the PCR mix was as follows (final volume of 20 uL):

| Reagent | Volume (uL) | Final concentration |
|---|---|---|
| $H_2O$ (milliQ) | 4 | - |
| 2x MyTaq | 10 | - |
| Forward primer (10 mM stock solution) | 2 | 1 µM |
| Reverse primer (10 mM stock solution) | 2 | 1µ |
| DNA sample (adjusted at 10 ng/µL) | 2 | 1 ng/uL |

Strain-specific primers used (all primers designed to have melting temperature of 60ºC):

| Primer | Sequence (5'-3') | Amplicon Size (bp) | Targeted Strain |
|---|---|---|---|
| PA7483_F2 | CCGGAACGTAAGCAACAT | 109* | Pa KABP021 (CECT7483) |
| PA7483_R2 | ACACCTCCATGATTTGATCC | | |
| LP7484_F3 | GCAATGTTTGCGGACTTAAA | 129 | Lp KABP022 (CECT7484) |
| LP7484_R3 | GCAACGTTCATAAGTTTCAAGA | | |
| LP7485_F1 | CCGTATTTGGCACGAGTT | 113 | Lp KABP023 (CECT7485) |
| LP7485_R1 | CTGTGCATACGTGGTTGT | | |
| LP33765_F3 | GGTTAAATTAAAGCCATAGAC | 242 | Lp KABP033 (CECT30292) |
| LP33765_R3 | TTAAGCAAGAAAAATGAGAAG | | |

*) An extra amplicon of 50bp can also be produced in this strain.

PCR products were separated and visualized in a 2% agarose gel using standard electrophoresis settings.

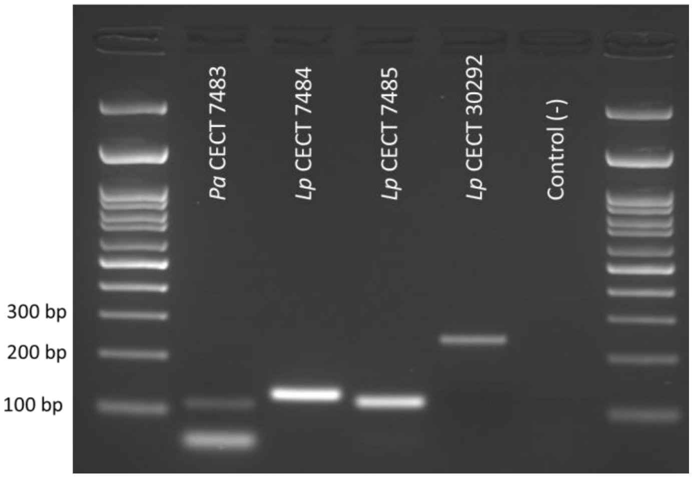

Representative image of an electrophoresis gel, each lane corresponding to a PCR reaction with specific pair of primers.

**Verification of Microbial Quality in Active Product and Placebo**

Active and placebo product batches were analysed after manufacture to comply the following specifications:

| **Target microorganism** | **Specification** | **Protocol** |
|---|---|---|
| Total yeasts and moulds | <10 cfu/gr | European Pharmacopoeia 2.6.12 |
| Bile-tolerant Gram-negatives | <10 cfu/gr | European Pharmacopoeia 2.6.13 |
| *Escherichia coli* | Absence in 1gr | |
| *Staphylococcus aureus* | Absence in 1gr | |
| *Salmonella* spp. | Absence in 25gr | |
| *Listeria monocytogenes* | Absence in 25gr | ISO11290-1 |

As a result of the testing, both the active and placebo product batches were found to comply with all of the above specifications.

#### Analytical Procedures in Nasopharyngeal and Serum Samples

SARS-CoV2-specific RT-qPCR

Nasopharynx swab samples for SARS-CoV2 RT-qPCR analysis were collected on each study site visit in 2.5 mL of Universal Transport Medium. Quantitative SARS-CoV2-specific RT-qPCR was performed using the Charité Berlin WHO protocol:[3]

Briefly, nucleic acids were first extracted from nasopharyngeal swabs in 2.5 mL of universal transport medium using RNA Minikit (Qiagen NV, Hilden, Germany); subsequently, RT-PCR was carried out in a QIAquant 96 5-plex instrument (Qiagen NV, Hilden, Germany) using primers RdRP_SARSr-F2 and RdRP_SARSr-R1, probe RdRP_SARSr-P2, and the Super Script III Platinum One Step qRT-PCR reagents (ThermoFisher, Waltham, US), using the following PCR mix (final volume of 25 microL):

| **Reagent** | **Volume (uL)** | **Final concentration** |
|---|---|---|
| $H_2O$ (RNAse free) | 2.1 | - |
| 2x Reaction Mix | 12.5 | - |
| MgSO4 (50mM) | 0.4 | 20 mM |
| Primer RdRp_SARSr-F2 (10 µM Stock solution) | 1.5 | 15 µM |
| Primer RdRp_SARSr-R1 (10 µM Stock solution) | 2.0 | 20 µM |
| Probe RdRp_SARSr-P2 (10 µM Stock solution) | 0.5 | 5 µM |
| SSIII/Taq EnzymeMix | 1.0 | - |
| Sample | 5.0 | Target |

Sequences of primers and of probe (5’-3’):

| | | |
|---|---|---|
| Forward | RdRP_SARSr-F2 | GTGARATGGTCATGTGTGGCGG |
| Reverse | RdRP_SARSr-R1 | CARATGTTAAASACACTATTAGCATA |
| Probe | RdRP_SARSr-P2 | FAM-CAGGTGGAACCTCATCAGGAGATGC-BBQ |

In order to translate cycle threshold (Ct) values into base 10 logarithm of viral copies/mL, each batch of RT-qPCR was reagents was calibrated using 10-fold dilutions of positive control plasmid containing the RdRP gene provided by Charité University of Medicine (Berlin, Germany), ranging $10^3$ to $10^9$ copies/mL of sample (dynamic range of the test). Negative results (*i.e.* <1 copy per reaction volume) were given a floor value of $10^2$ copies/mL.

Other Assays

10 mL of venous blood samples were taken on each study visit and stored at -70ºC until the end of the study for analysis. Assays for SARS-CoV2 spike protein-specific IgG (DiaSorin SpA, Saluggia, Italy), SARS-CoV2 spike protein-specific IgM (Abbot Laboratories, IL, US), D-dimer (Spinreact SA, Sant Esteve de Bas, Spain) and hsCRP (Abbot Laboratories, IL, US) were performed on plasma with EDTA, according to manufacturers’ instructions and using manufacturer’s standards for calibration curves.

Assay Laboratory

All these analytical procedures were conducted at DiagnoMol Laboratories (Mexico City, Mexico), and methodology details. The laboratory is ISO15189-accredited (requirements for quality and competence in laboratories analyzing medical samples of human origin).

#### Supplementary Results

**Figure S1. Serum levels of high-sensitivity C-Reactive Protein (hsCRP) and D-Dimer.**

Geometric means of serum levels of high-sensitivity hs-CRP (A) and of D-Dimer (B) across the study. Error bars denote 95%CI of the geometric means.

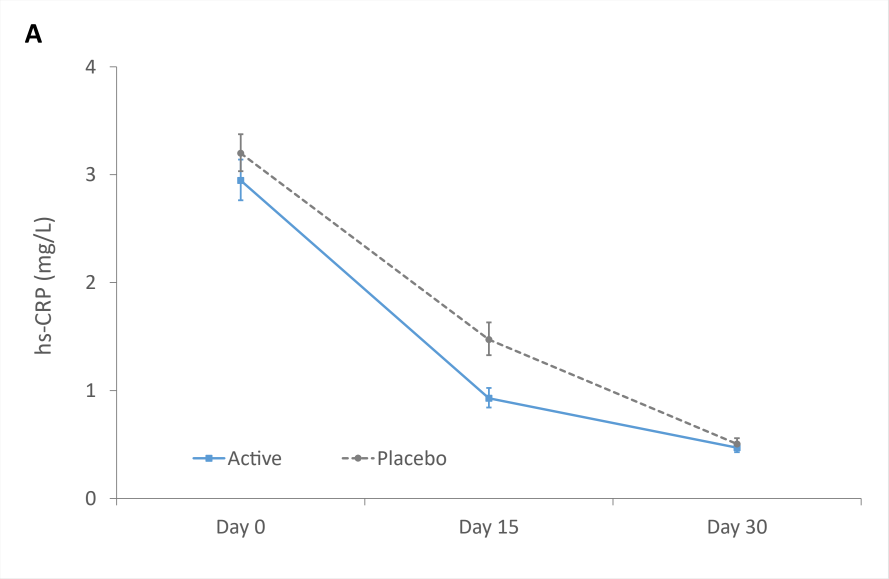

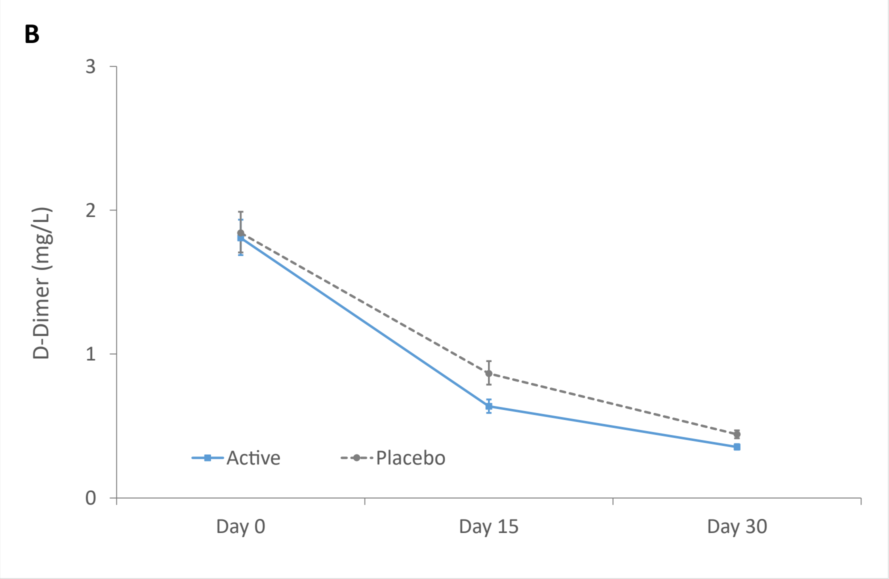

| Participants | Day 0 | Day 15 | Day 30 |
|---|---|---|---|
| • Active (n) | 150 | 148 | 147 |
| • Placebo (n) | 150 | 149 | 146 |

**Figure S2. Rating of lung abnormalities.**

Means values of lung abnormality rating according to the Brixia score. Error bars denote 95%CI of the means.

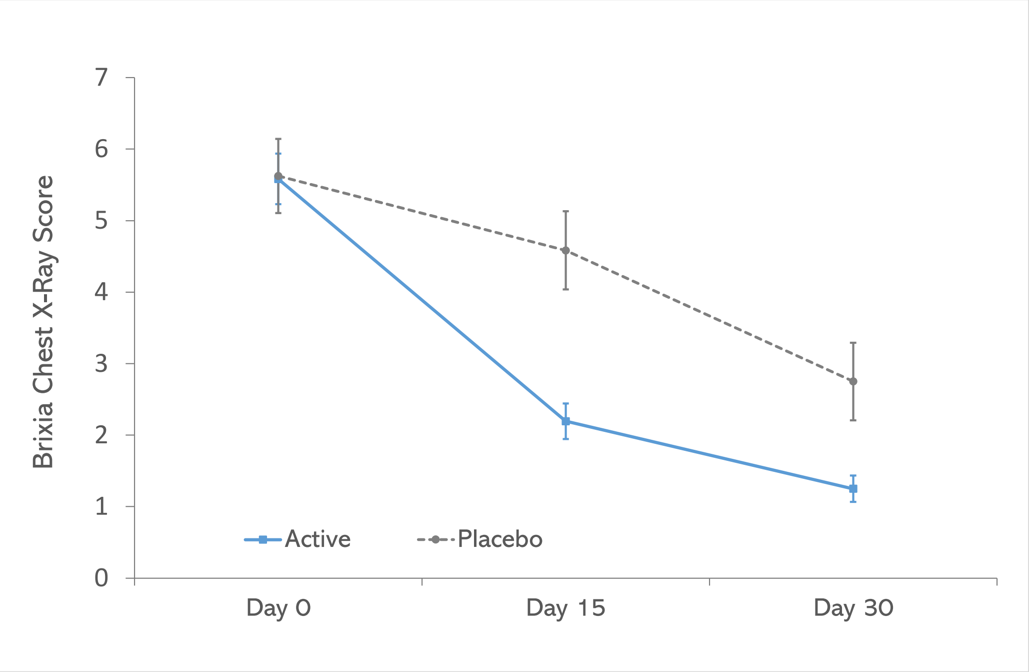

| Participants | Day 0 | Day 15 | Day 30 |
|---|---|---|---|
| • Active (n) | 72 | 72 | 72 |
| • Placebo (n) | 48 | 48 | 48 |

**Figure S3. Post-hoc analysis of time to overall symptom-resolution, according to participants diaries.**

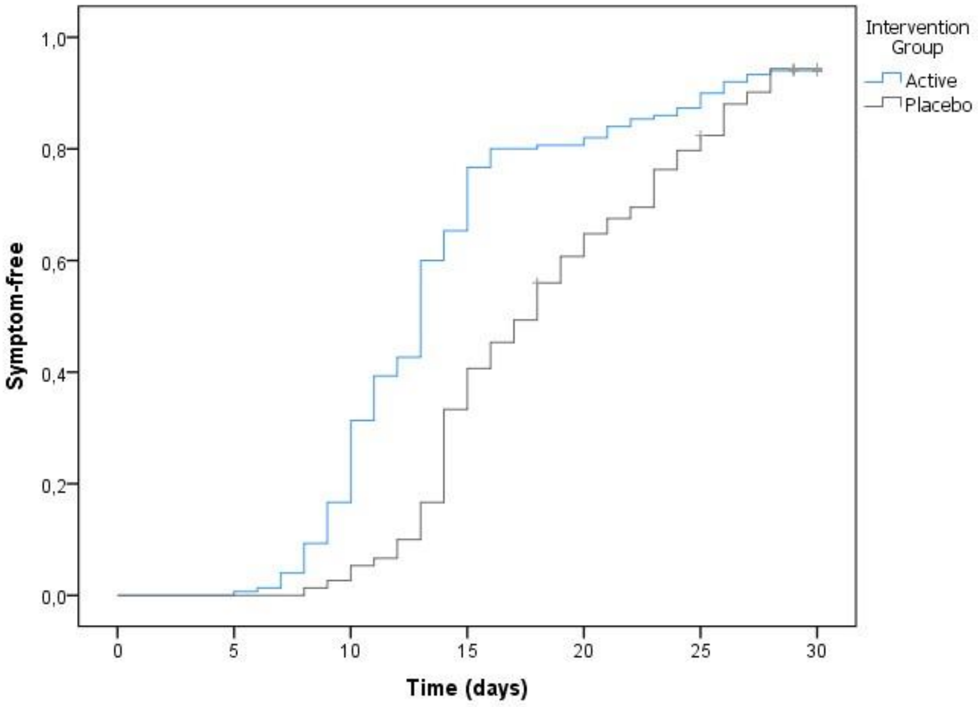

Overall symptom resolution was based on the five symptoms key considered in the study: fever (>37.5ºC), cough, headache, shortness of breath and body aches.

Symptomatic subjects remaining (number at risk) and censored subjects per study group:

| | D 0 | D 3 | D 6 | D 9 | D 12 | D 15 | D 18 | D 21 | D 24 | D 27 | D 30 |
|---|---|---|---|---|---|---|---|---|---|---|---|
| **Active** | 150 | 150 | 149 | 136 | 91 | 52 | 30 | 27 | 21 | 12 | NA |
| Censored | 0 | 0 | 0 | 0 | 0 | 0 | 0 | 0 | 0 | 0 | 9 |
| **Placebo** | 150 | 150 | 150 | 148 | 140 | 100 | 76 | 52 | 35 | 17 | NA |
| Censored | 0 | 0 | 0 | 0 | 0 | 0 | 1 | 0 | 1 | 0 | 8 |

(*) Subjects who were not negative for symptoms for at least 24h at study exit were censored.

Kaplan-Meyer estimates:

| | Mean duration (days) | 95% CI | Median duration (days) | 95% CI |
|---|---|---|---|---|
| **Active** | 14.4 | 13.4 – 15.4 | 13.0 | 12.5 – 13.5 |
| **Placebo** | 18.6 | 17.7 – 19.6 | 18.0 | 16.4 – 19.6 |

**Table S1. Remission status of study subjects at the end of the study (visit 3).**

| Group | RT-qPCR and Symptomatic Status | Classification |
|---|---|---|
| Active (n=147) | 82 subjects had a negative RT-qPCR on visit 3, of which: | |
| | • Subject #55 had 4 days of fever (days 27-30) | Non-remitter |
| | • Subjects #106 and #134 had 5 days of fever (days 25-29) | Non-remitter |
| | • Subject #139 had 5 days of fever (days 26-30) | Non-remitter |
| | • Subject #163 had isolated fever on last day, but symptoms had stopped on day 11, and had no fever since day 0 | Adverse Event & Remitter |
| | • Subject #214 had isolated fever on last day, but symptoms had stopped on day 5, and had no fever since day 2 | Adverse Event & Remitter |
| | • The remaining 76 subjects were asymptomatic for at least 24h before the last visit | Remitters |
| | Therefore, 78 subjects were considered as remitters (53.1%) | |
| Placebo (n=146) | 41 subjects had a negative RT-qPCR on visit 3, of which: | |
| | • Subject #249 had isolated fever on day 30, but symptoms had stopped on day 14, had negative RT-qPCR on visit 2 and had no fever since day 4 | Adverse Event & Remitter |
| | • The remaining 40 subjects were asymptomatic for at least 24h before the last visit | Remitters |
| | Therefore, 41 subjects were considered as remitters (28.1%) | |

**Table S2. Post-hoc sensitivity analysis on remission status (visit 3): logistic regression before and after adjusting for baseline imbalances.**

1) Unadjusted logistic regression of remission vs. study group (n = 293):

| -2 log likelihood | Cox & Snell $R^2$ | Nagelkerke $R^2$ |
|---|---|---|
| 376.602 | 0.063 | 0.086 |

| | B | Wald | df | P-value | OR | 95%CI of OR | |
|---|---|---|---|---|---|---|---|
| Study group (active) | 1.063 | 18.455 | 1 | <0.001 | 2.895 | 1.782 | 4.702 |
| Constant | -0.940 | 26.076 | 1 | <0.001 | - | - | - |

2) Adjusted logistic regression of remission vs. study group, Body Mass Index (BMI), shortness of breath (dyspnea), peripheral oxygen saturation ($SpO_2$) and lung abnormalities as per chest X-ray imaging (n = 293):

| -2 log likelihood | Cox & Snell $R^2$ | Nagelkerke $R^2$ |
|---|---|---|
| 371.485 | 0.080 | 0.107 |

| | B | Wald | df | P-value | OR | 95%CI of OR | |
|---|---|---|---|---|---|---|---|
| Study group (active) | 1.092 | 16.731 | 1 | <0.001 | 2.982 | 1.766 | 5.032 |
| BMI at baseline (Kg/m2) | -0.001 | 0.000 | 1 | 0.984 | 0.999 | 0.947 | 1.055 |
| Dyspnea at baseline (yes) | -0.459 | 2.934 | 1 | 0.087 | 0.632 | 0.374 | 1.068 |
| SpO2 at baseline (%) | -0.186 | 1.688 | 1 | 0.194 | 0.830 | 0.627 | 1.099 |
| Lung abnormalities (yes) | -0.033 | 0.016 | 1 | 0.900 | 0.968 | 0.583 | 1.607 |
| Constant | 16.255 | 1.567 | 1 | 0.211 | - | - | - |

**Table S3. Post-hoc analyses on remission status across different study subpopulations at the end of the study (visit 3).**

| Subpopulation | Active | Placebo | P-value |
|---|---|---|---|
| Age | | | |
| • Less than 50 years old (n=235) | 63 of 124 (50.8%) | 31 of 111 (27.9%) | 0.0004 |
| • 50 years and older (n=58) | 15 of 23 (65.2%) | 10 of 35 (28.6%) | 0.0063 |
| Sex | | | |
| • Female (n=155) | 45 of 79 (57.0%) | 25 of 76 (32.9%) | 0.0027 |
| • Male (n=138) | 33 of 68 (48.5%) | 16 of 70 (22.9%) | 0.0017 |
| Viral load at baseline | | | |
| • Viral load ≤ baseline median (n=152) | 55 of 83 (66.3%) | 27 of 69 (39.1%) | 0.0009 |
| • Viral load > baseline median (n=141) | 23 of 64 (35.9%) | 14 of 77 (18.2%) | 0.0174 |
| Metabolic risk factors | | | |
| • Diabetes, hypertension or BMI ≥30 (n=121) | 29 of 53 (54.7%) | 19 of 68 (27.9%) | 0.0029 |
| • None of the above (n=172) | 49 of 94 (52.1%) | 22 of 78 (28.2%) | 0.0016 |
| Time from symptom onset | | | |
| • One to four days (n=200) | 55 of 103 (53.4%) | 29 of 97 (29.9%) | 0.0008 |
| • Five to seven days (n=93) | 23 of 44 (52.3%) | 12 of 49 (24.5%) | 0.0060 |

**Table S4. Post-hoc analyses on duration of acetaminophen use, loss of taste and smell.**

Duration, split by baseline status for each symptom (yes/no), indicated as median days [range]. Number of subjects in each subgroup are indicated within parentheses below.

| Symptom | Baseline status | Active | Placebo | P-value |
|---|---|---|---|---|
| Loss of taste | Yes | 3 [1-7] (n=55) | 7 [2-13] (n=62) | < 0.0001 |
| | No | 0 [0-4] (n=95) | 0 [0-6] (n=88) | 0.2109 |
| Loss of smell | Yes | 3 [1-12] (n=57) | 8 [3-14] (n=66) | < 0.0001 |
| | No | 0 [0-4] (n=93) | 0 [0-0] (n=84) | 0.0978 |
| Acetaminophen use | Yes | 1 [0-12] (n=83) | 3 [1-14] (n=70) | < 0.0001 |
| | No | 1 [0-10] (n=67) | 3 [1-15] (n=80) | < 0.0001 |

**Table S5. Presence of the *plnG-plnH* gene cluster.**

Presence of the p*lnG-plnH* gene cluster in the genomes of the *L. plantarum* strains used in this study. This gene cluster is known be recognized by various types of antigen-presenting cells:[1,2]

| **Bacterial Strain** | **Presence of *plnG-plnH* *** |
|---|---|
| *L. plantarum* KABP022 | Yes, 1 genomic copy (99% identity) |
| *L. plantarum* KABP023 | Yes, 1 genomic copy (99% identity) |
| *L. plantarum* KABP033 | Yes, 1 genomic and plasmidic copies (all 99% identity) |

(*) Presence was determined as follows: Sequences of genes *plnG* and *plnH* (loci lp_0423 and lp_0424, respectively) were downloaded from the reference *Lactiplantibacillus plantarum* genome in GenBank (strain WCSF1; https://www.ncbi.nlm.nih.gov/nuccore/AL935263) and searched against the complete genomes of strains KABP022, KABP023 and KABP033, using BLASTn program[3] and a coverage threshold of >95%.

### Annex: CONSORT 2010 Checklist

#### CONSORT 2010 checklist of information to include when reporting a randomised trial*

| Section/Topic | Item No | Checklist item | Reported on page No |
|---|---|---|---|
| **Title and abstract** | | | |
| | 1a | Identification as a randomised trial in the title | 1 |
| | 1b | Structured summary of trial design, methods, results, and conclusions (for specific guidance see CONSORT for abstracts) | 1 |
| **Introduction** | | | |
| Background and objectives | 2a | Scientific background and explanation of rationale | 2 - 3 |
| | 2b | Specific objectives or hypotheses | 3 |
| **Methods** | | | |
| Trial design | 3a | Description of trial design (such as parallel, factorial) including allocation ratio | 4 |
| | 3b | Important changes to methods after trial commencement (such as eligibility criteria), with reasons | not applicable |
| Participants | 4a | Eligibility criteria for participants | 4, Suppl. Methods |
| | 4b | Settings and locations where the data were collected | 4 |
| Interventions | 5 | The interventions for each group with sufficient details to allow replication, including how and when they were actually administered | 5 - 6 |
| Outcomes | 6a | Completely defined pre-specified primary and secondary outcome measures, including how and when they were assessed | 5 - 7 |
| | 6b | Any changes to trial outcomes after the trial commenced, with reasons | 6 - 7 |
| Sample size | 7a | How sample size was determined | 7 - 8 |
| | 7b | When applicable, explanation of any interim analyses and stopping guidelines | not applicable |
| Randomisation: | | | |
| Sequence generation | 8a | Method used to generate the random allocation sequence | 4 - 5 |
| | 8b | Type of randomisation; details of any restriction (such as blocking and block size) | 4 |
| Allocation concealment mechanism | 9 | Mechanism used to implement the random allocation sequence (such as sequentially numbered containers), describing any steps taken to conceal the sequence until interventions were assigned | 4 |
| Implementation | 10 | Who generated the random allocation sequence, who enrolled participants, and who assigned participants to interventions | 4 |
| Blinding | 11a | If done, who was blinded after assignment to interventions (for example, participants, care providers, those | |

| | | | |
|---|---|---|---|
| | | assessing outcomes) and how | 4 |
| | 11b | If relevant, description of the similarity of interventions | 5 |
| Statistical methods | 12a | Statistical methods used to compare groups for primary and secondary outcomes | 8 |
| | 12b | Methods for additional analyses, such as subgroup analyses and adjusted analyses | 8 |
| **Results** | | | |
| Participant flow (a diagram is strongly recommended) | 13a | For each group, the numbers of participants who were randomly assigned, received intended treatment, and were analysed for the primary outcome | 9, Fig.1 |
| | 13b | For each group, losses and exclusions after randomisation, together with reasons | 19, Fig.1 |
| Recruitment | 14a | Dates defining the periods of recruitment and follow-up | 9 |
| | 14b | Why the trial ended or was stopped | not applicable |
| Baseline data | 15 | A table showing baseline demographic and clinical characteristics for each group | Table 1 |
| Numbers analysed | 16 | For each group, number of participants (denominator) included in each analysis and whether the analysis was by original assigned groups | 8; Fig. 1 |
| Outcomes and estimation | 17a | For each primary and secondary outcome, results for each group, and the estimated effect size and its precision (such as 95% confidence interval) | Table 2; Fig.2,S1,S2 |
| | 17b | For binary outcomes, presentation of both absolute and relative effect sizes is recommended | 9 |
| Ancillary analyses | 18 | Results of any other analyses performed, including subgroup analyses and adjusted analyses, distinguishing pre-specified from exploratory | 10; Fig.S3; Table S2-S4 |
| Harms | 19 | All important harms or unintended effects in each group (for specific guidance see CONSORT for harms) | 10; Table 3 |
| **Discussion** | | | |
| Limitations | 20 | Trial limitations, addressing sources of potential bias, imprecision, and, if relevant, multiplicity of analyses | 11,13 |
| Generalisability | 21 | Generalisability (external validity, applicability) of the trial findings | 11,13 |
| Interpretation | 22 | Interpretation consistent with results, balancing benefits and harms, and considering other relevant evidence | 14 |
| **Other information** | | | |
| Registration | 23 | Registration number and name of trial registry | 1,4 |
| Protocol | 24 | Where the full trial protocol can be accessed, if available | yes, online |
| Funding | 25 | Sources of funding and other support (such as supply of drugs), role of funders | 14,15 |

*We strongly recommend reading this statement in conjunction with the CONSORT 2010 Explanation and Elaboration for important clarifications on all the items. If relevant, we also recommend reading CONSORT extensions for cluster randomised trials, non-inferiority and equivalence trials, non-pharmacological treatments, herbal interventions, and pragmatic trials. Additional extensions are forthcoming: for those and for up to date references relevant to this checklist, see www.consort-statement.org.
