## Supplementary material for "Efficacy and safety of novel probiotic formulation in adult Covid19 outpatients: a randomized, placebo-controlled clinical trial": ICMJE Conflict of Interest Statements

### ICMJE DISCLOSURE FORM

**Date: May, 10th, 2021**
**Your Name: Ana Teresa Abreu Abreu**
**Manuscript Title: Efficacy and safety of probiotic formulation in adult COVID19 outpatients: a randomized, placebo-controlled clinical trial**
**Manuscript number (if known): N.A.**

**In the interest of transparency, we ask you to disclose all relationships/activities/interests listed below that are related to the content of your manuscript. "Related" means any relation with for-profit or not-for-profit third parties whose interests may be affected by the content of the manuscript. Disclosure represents a commitment to transparency and does not necessarily indicate a bias. If you are in doubt about whether to list a relationship/activity/interest, it is preferable that you do so.**

**The following questions apply to the author's relationships/activities/interests as they relate to the current manuscript only.**

**The author's relationships/activities/interests should be defined broadly. For example, if your manuscript pertains to the epidemiology of hypertension, you should declare all relationships with manufacturers of antihypertensive medication, even if that medication is not mentioned in the manuscript.**

**In item #1 below, report all support for the work reported in this manuscript without time limit. For all other items, the time frame for disclosure is the past 36 months.**

| | | **Name all entities with whom you have this relationship or indicate none (add rows as needed)** | **Specifications/Comments (e.g., if payments were made to you or to your institution)** |
|---|---|---|---|
| **Time frame: Since the initial planning of the work** | | | |
| 1 | All support for the present manuscript (e.g., funding, provision of study materials, medical writing, article processing charges, etc.) **No time limit for this item.** | **AB-BIOTICS SA** | Innovación y Desarrollo de Estrategias en Salud SAdeCV which leader the Project received the financial support from AB-BIOTCIS that help to cover the activities related to the develop of the different activities where I participated |
| **Time frame: past 36 months** | | | |
| 2 | Grants or contracts from any entity (if not indicated in item #1 above). | None | |
| 3 | Royalties or licenses | None | |
| 4 | Consulting fees | None | |

| | | | |
|---|---|---|---|
| 5 | Payment or honoraria for lectures, presentations, speakers bureaus, manuscript writing or educational events | AB-BIOTICS | Participating as speaker |
| | | BioGaia | Participating as speaker |
| | | Mayoly-Splinder | Participating as speaker |
| 6 | Payment for expert testimony | None | |
| 7 | Support for attending meetings and/or travel | None | |
| 8 | Patents planned, issued or pending | None | |
| 9 | Participation on a Data Safety Monitoring Board or Advisory Board | None | |
| 10 | Leadership or fiduciary role in other board, society, committee or advocacy group, paid or unpaid | None | |
| 11 | Stock or stock options | None | |
| 12 | Receipt of equipment, materials, drugs, medical writing, gifts or other services | None | |
| 13 | Other financial or non-financial interests | None | |

**Please place an "X" next to the following statement to indicate your agreement:**

**X I certify that I have answered every question and have not altered the wording of any of the questions on this form.**

**Dra. Ana Teresa Abreu-Abreu**

#### ICMJE DISCLOSURE FORM

Date: May, 10th, 2021
Your Name: Cesar D. Nieto-Rufino
Manuscript Title: Efficacy and safety of probiotic formulation in adult Covid19 outpatients: a randomized, placebo-controlled clinical trial
Manuscript number (if known): N.A.

In the interest of transparency, we ask you to disclose all relationships/activities/interests listed below that are related to the content of your manuscript. "Related" means any relation with for-profit or not-for-profit third parties whose interests may be affected by the content of the manuscript. Disclosure represents a commitment to transparency and does not necessarily indicate a bias. If you are in doubt about whether to list a relationship/activity/interest, it is preferable that you do so.

The following questions apply to the author's relationships/activities/interests as they relate to the current manuscript only.

The author's relationships/activities/interests should be defined broadly. For example, if your manuscript pertains to the epidemiology of hypertension, you should declare all relationships with manufacturers of antihypertensive medication, even if that medication is not mentioned in the manuscript.

In item #1 below, report all support for the work reported in this manuscript without time limit. For all other items, the time frame for disclosure is the past 36 months.

| | | Name all entities with whom you have this relationship or indicate none (add rows as needed) | Specifications/Comments (e.g., if payments were made to you or to your institution) |
|---|---|---|---|
| | | **Time frame: Since the initial planning of the work** | |
| 1 | All support for the present manuscript (e.g., funding, provision of study materials, medical writing, article processing charges, etc.) **No time limit for this item.** | AB-BIOTICS SA | Innovación y Desarrollo de Estrategias en Salud SAdeCV who leader the Project received the financial support from AB-BIOTCIS that help to cover the activities related to the develop of the different activities where I participated |
| | | **Time frame: past 36 months** | |
| 2 | Grants or contracts from any entity (if not indicated in item #1 above). | None | |
| 3 | Royalties or licenses | None | |
| 4 | Consulting fees | None | |

| | | |
|---|---|---|
| 5 | Payment or honoraria for lectures, presentations, speakers bureaus, manuscript writing or educational events | None |
| 6 | Payment for expert testimony | None |
| 7 | Support for attending meetings and/or travel | None |
| 8 | Patents planned, issued or pending | None |
| 9 | Participation on a Data Safety Monitoring Board or Advisory Board | None |
| 10 | Leadership or fiduciary role in other board, society, committee or advocacy group, paid or unpaid | None |
| 11 | Stock or stock options | None |
| 12 | Receipt of equipment, materials, drugs, medical writing, gifts or other services | None |
| 13 | Other financial or non-financial interests | None |

**Please place an "X" next to the following statement to indicate your agreement:**

**X I certify thatI have answered every question and have not altered the wording of any of the questions on this form.**

**Dr. Cesar D. Nieto-Rufino**

#### ICMJE DISCLOSURE FORM

**Date: May, 10th, 2021**
**Your Name: Carlos Jiménez Gutiérrez**
**Manuscript Title: Efficacy and safety of probiotic formulation in adult COVID19 outpatients: a randomized, placebo-controlled clinical trial**
**Manuscript number (if known): N.A.**

**In the interest of transparency, we ask you to disclose all relationships/activities/interests listed below that are related to the content of your manuscript. "Related" means any relation with for-profit or not-for-profit third parties whose interests may be affected by the content of the manuscript. Disclosure represents a commitment to transparency and does not necessarily indicate a bias. If you are in doubt about whether to list a relationship/activity/interest, it is preferable that you do so.**

**The following questions apply to the author's relationships/activities/interests as they relate to the current manuscript only.**

**The author's relationships/activities/interests should be defined broadly. For example, if your manuscript pertains to the epidemiology of hypertension, you should declare all relationships with manufacturers of antihypertensive medication, even if that medication is not mentioned in the manuscript.**

**In item #1 below, report all support for the work reported in this manuscript without time limit. For all other items, the time frame for disclosure is the past 36 months.**

| | | Name all entities with whom you have this relationship or indicate none (add rows as needed) | Specifications/Comments (e.g., if payments were made to you or to your institution) |
|---|---|---|---|
| | | **Time frame: Since the initial planning of the work** | |
| 1 | All support for the present manuscript (e.g., funding, provision of study materials, medical writing, article processing charges, etc.) **No time limit for this item.** | AB BIOTICS S.A. | Innovación y Desarrollo de Estrategias en Salud SAdeCV which leader the Project received the financial support from AB-BIOTCIS that help to cover the activities related to the develop of the different activities where I participated |
| | | **Time frame: past 36 months** | |
| 2 | Grants or contracts from any entity (if not indicated in item #1 above). | None | |
| 3 | Royalties or licenses | None | |
| 4 | Consulting fees | None | |

| | | |
|---|---|---|
| 5 | Payment or honoraria for lectures, presentations, speakers bureaus, manuscript writing or educational events | None |
| 6 | Payment for expert testimony | None |
| 7 | Support for attending meetings and/or travel | None |
| 8 | Patents planned, issued or pending | None |
| 9 | Participation on a Data Safety Monitoring Board or Advisory Board | None |
| 10 | Leadership or fiduciary role in other board, society, committee or advocacy group, paid or unpaid | None |
| 11 | Stock or stock options | None |
| 12 | Receipt of equipment, materials, drugs, medical writing, gifts or other services | None |
| 13 | Other financial or non-financial interests | None |

**Please place an "X" next to the following statement to indicate your agreement:**

**X I certify that I have answered every question and have not altered the wording of any of the questions on this form.**

**Dr. Carlos Jiménez-Gutiérrez**

#### ICMJE DISCLOSURE FORM

**Date: May, 10th, 2021**
**Your Name: Eduardo López-Orduña**
**Manuscript Title: Efficacy and safety of probiotic formulation in adult COVID19 outpatients: a randomized, placebo-controlled clinical trial**
**Manuscript number (if known): N.A.**

**In the interest of transparency, we ask you to disclose all relationships/activities/interests listed below that are related to the content of your manuscript. "Related" means any relation with for-profit or not-for-profit third parties whose interests may be affected by the content of the manuscript. Disclosure represents a commitment to transparency and does not necessarily indicate a bias. If you are in doubt about whether to list a relationship/activity/interest, it is preferable that you do so.**

**The following questions apply to the author's relationships/activities/interests as they relate to the current manuscript only.**

**The author's relationships/activities/interests should be defined broadly. For example, if your manuscript pertains to the epidemiology of hypertension, you should declare all relationships with manufacturers of antihypertensive medication, even if that medication is not mentioned in the manuscript.**

**In item #1 below, report all support for the work reported in this manuscript without time limit. For all other items, the time frame for disclosure is the past 36 months.**

| | | **Name all entities with whom you have this relationship or indicate none (add rows as needed)** | **Specifications/Comments (e.g., if payments were made to you or to your institution)** |
|---|---|---|---|
| | | **Time frame: Since the initial planning of the work** | |
| 1 | All support for the present manuscript (e.g., funding, provision of study materials, medical writing, article processing charges, etc.) **No time limit for this item.** | AB-BIOTICS SA | Innovación y Desarrollo de Estrategias en Salud SAdeCV who leader the Project received the financial support from AB-BIOTCIS that help to cover the activities related to the develop of the different activities where I participated |
| | | **Time frame: past 36 months** | |
| 2 | Grants or contracts from any entity (if not indicated in item #1 above). | None | |
| 3 | Royalties or licenses | None | |
| 4 | Consulting fees | None | |

| | | |
|---|---|---|
| 5 | Payment or honoraria for lectures, presentations, speakers bureaus, manuscript writing or educational events | None |
| 6 | Payment for expert testimony | None |
| 7 | Support for attending meetings and/or travel | None |
| 8 | Patents planned, issued or pending | None |
| 9 | Participation on a Data Safety Monitoring Board or Advisory Board | None |
| 10 | Leadership or fiduciary role in other board, society, committee or advocacy group, paid or unpaid | None |
| 11 | Stock or stock options | None |
| 12 | Receipt of equipment, materials, drugs, medical writing, gifts or other services | None |
| 13 | Other financial or non-financial interests | None |

**Please place an "X" next to the following statement to indicate your agreement:**

**X I certify that I have answered every question and have not altered the wording of any of the questions on this form.**

X

Eduardo López Orduña

### ICMJE DISCLOSURE FORM

**Date: May, 10th, 2021**
**Your Name: Gabriel López-Velázquez**
**Manuscript Title: Efficacy and safety of probiotic formulation in adult COVID19 outpatients: a randomized, placebo-controlled clinical trial**
**Manuscript number (if known): N.A.**

**In the interest of transparency, we ask you to disclose all relationships/activities/interests listed below that are related to the content of your manuscript. "Related" means any relation with for-profit or not-for-profit third parties whose interests may be affected by the content of the manuscript. Disclosure represents a commitment to transparency and does not necessarily indicate a bias. If you are in doubt about whether to list a relationship/activity/interest, it is preferable that you do so.**

**The following questions apply to the author's relationships/activities/interests as they relate to the current manuscript only.**

**The author's relationships/activities/interests should be defined broadly. For example, if your manuscript pertains to the epidemiology of hypertension, you should declare all relationships with manufacturers of antihypertensive medication, even if that medication is not mentioned in the manuscript.**

**In item #1 below, report all support for the work reported in this manuscript without time limit. For all other items, the time frame for disclosure is the past 36 months.**

| | | **Name all entities with whom you have this relationship or indicate none (add rows as needed)** | **Specifications/Comments (e.g., if payments were made to you or to your institution)** |
|---|---|---|---|
| | | **Time frame: Since the initial planning of the work** | |
| 1 | All support for the present manuscript (e.g., funding, provision of study materials, medical writing, article processing charges, etc.) **No time limit for this item.** | AB-BIOTICS SA | Innovación y Desarrollo de Estrategias en Salud SAdeCV which leader the Project received the financial support from AB-BIOTCIS that help to cover the activities related to the develop of the different activities where I participated |
| | | **Time frame: past 36 months** | |
| 2 | Grants or contracts from any entity (if not indicated in item #1 above). | None | |
| 3 | Royalties or licenses | None | |
| 4 | Consulting fees | None | |

| | | |
|---|---|---|
| 5 | Payment or honoraria for lectures, presentations, speakers bureaus, manuscript writing or educational events | None |
| 6 | Payment for expert testimony | None |
| 7 | Support for attending meetings and/or travel | None |
| 8 | Patents planned, issued or pending | None |
| 9 | Participation on a Data Safety Monitoring Board or Advisory Board | None |
| 10 | Leadership or fiduciary role in other board, society, committee or advocacy group, paid or unpaid | None |
| 11 | Stock or stock options | None |
| 12 | Receipt of equipment, materials, drugs, medical writing, gifts or other services | None |
| 13 | Other financial or non-financial interests | None |

**Please place an "X" next to the following statement to indicate your agreement:**

**X I certify that I have answered every question and have not altered the wording of any of the questions on this form.**

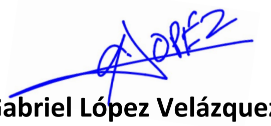

**Dr. Gabriel López Velázquez**

#### ICMJE DISCLOSURE FORM

**Date: May, 10th, 2021**
**Your Name: Irma Jiménez-Escobar**
**Manuscript Title: Efficacy and safety of probiotic formulation in adult COVID19 outpatients: a randomized, placebo-controlled clinical trial**
**Manuscript number (if known): N.A.**

**In the interest of transparency, we ask you to disclose all relationships/activities/interests listed below that are related to the content of your manuscript. "Related" means any relation with for-profit or not-for-profit third parties whose interests may be affected by the content of the manuscript. Disclosure represents a commitment to transparency and does not necessarily indicate a bias. If you are in doubt about whether to list a relationship/activity/interest, it is preferable that you do so.**

**The following questions apply to the author's relationships/activities/interests as they relate to the current manuscript only.**

**The author's relationships/activities/interests should be defined broadly. For example, if your manuscript pertains to the epidemiology of hypertension, you should declare all relationships with manufacturers of antihypertensive medication, even if that medication is not mentioned in the manuscript.**

**In item #1 below, report all support for the work reported in this manuscript without time limit. For all other items, the time frame for disclosure is the past 36 months.**

| | | **Name all entities with whom you have this relationship or indicate none (add rows as needed)** | **Specifications/Comments (e.g., if payments were made to you or to your institution)** |
|---|---|---|---|
| **Time frame: Since the initial planning of the work** | | | |
| 1 | All support for the present manuscript (e.g., funding, provision of study materials, medical writing, article processing charges, etc.) **No time limit for this item.** | AB-BIOTICS SA | Innovación y Desarrollo de Estrategias en Salud SAdeCV which leader the Project received the financial support from AB-BIOTCIS that help to cover the activities related to the develop of the different activities where I participated |
| **Time frame: past 36 months** | | | |
| 2 | Grants or contracts from any entity (if not indicated in item #1 above). | None | |
| 3 | Royalties or licenses | None | |
| 4 | Consulting fees | None | |

| | | |
|---|---|---|
| 5 | Payment or honoraria for lectures, presentations, speakers bureaus, manuscript writing or educational events | None |
| 6 | Payment for expert testimony | None |
| 7 | Support for attending meetings and/or travel | None |
| 8 | Patents planned, issued or pending | None |
| 9 | Participation on a Data Safety Monitoring Board or Advisory Board | None |
| 10 | Leadership or fiduciary role in other board, society, committee or advocacy group, paid or unpaid | None |
| 11 | Stock or stock options | None |
| 12 | Receipt of equipment, materials, drugs, medical writing, gifts or other services | None |
| 13 | Other financial or non-financial interests | None |

**Please place an "X" next to the following statement to indicate your agreement:**

**X I certify that I have answered every question and have not altered the wording of any of the questions on this form.**

Irma Jimenez-Escobar MD, MHM

#### ICMJE DISCLOSURE FORM

**Date:** April 30th, 2021
**Your Name:** Jordi Espadaler Mazo
**Manuscript Title:** Efficacy and safety of probiotic formulation in adult Covid19 outpatients: a randomized, placebo-controlled clinical trial
**Manuscript number (if known):** N.A

**In the interest of transparency, we ask you to disclose all relationships/activities/interests listed below that are related to the content of your manuscript. "Related" means any relation with for-profit or not-for-profit third parties whose interests may be affected by the content of the manuscript. Disclosure represents a commitment to transparency and does not necessarily indicate a bias. If you are in doubt about whether to list a relationship/activity/interest, it is preferable that you do so.**

**The following questions apply to the author's relationships/activities/interests as they relate to the current manuscript only.**

**The author's relationships/activities/interests should be defined broadly. For example, if your manuscript pertains to the epidemiology of hypertension, you should declare all relationships with manufacturers of antihypertensive medication, even if that medication is not mentioned in the manuscript.**

**In item #1 below, report all support for the work reported in this manuscript without time limit. For all other items, the time frame for disclosure is the past 36 months.**

| | | **Name all entities with whom you have this relationship or indicate none (add rows as needed)** | **Specifications/Comments (e.g., if payments were made to you or to your institution)** |
|---|---|---|---|
| | | Time frame: Since the initial planning of the work | |
| 1 | All support for the present manuscript (e.g., funding, provision of study materials, medical writing, article processing charges, etc.) **No time limit for this item.** | Kaneka AB-BIOTICS S.A. | Full-time employee at Kaneka AB-BIOTICS S.A. (Spain) |
| | | Time frame: past 36 months | |
| 2 | Grants or contracts from any entity (if not indicated in item #1 above). | ____None | |
| 3 | Royalties or licenses | ____None | |
| 4 | Consulting fees | ____None | |

| | | | |
|---|---|---|---|
| 5 | Payment or honoraria for lectures, presentations, speakers bureaus, manuscript writing or educational events | ____None | |
| 6 | Payment for expert testimony | ____None | |
| 7 | Support for attending meetings and/or travel | ____None | |
| 8 | Patents planned, issued or pending | Patent pending on strain KABP033 (CECT30292) | Featured as inventor (but not patent applicant) of patent "PROBIOTIC COMPOSITION FOR THE TREATMENT OF COVID-19", PCT/EP2021/053539 |
| 9 | Participation on a Data Safety Monitoring Board or Advisory Board | ____None | |
| 10 | Leadership or fiduciary role in other board, society, committee or advocacy group, paid or unpaid | ____None | |
| 11 | Stock or stock options | ____None | |
| 12 | Receipt of equipment, materials, drugs, medical writing, gifts or other services | ____None | |
| 13 | Other financial or non-financial interests | ____None | |

**Please place an "X" next to the following statement to indicate your agreement:**

X **I certify that I have answered every question and have not altered the wording of any of the questions on this form.**

### ICMJE DISCLOSURE FORM

**Date: Apr, 30th, 2021**
**Your Name: Pedro Gutierrez Castrellon**
**Manuscript Title: Efficacy and safety of probiotic formulation in adult Covid19 outpatients: a randomized, placebo-controlled clinical trial**
**Manuscript number (if known):** NA

**In the interest of transparency, we ask you to disclose all relationships/activities/interests listed below that are related to the content of your manuscript. "Related" means any relation with for-profit or not-for-profit third parties whose interests may be affected by the content of the manuscript. Disclosure represents a commitment to transparency and does not necessarily indicate a bias. If you are in doubt about whether to list a relationship/activity/interest, it is preferable that you do so.**

**The following questions apply to the author's relationships/activities/interests as they relate to the current manuscript only.**

**The author's relationships/activities/interests should be defined broadly. For example, if your manuscript pertains to the epidemiology of hypertension, you should declare all relationships with manufacturers of antihypertensive medication, even if that medication is not mentioned in the manuscript.**

**In item #1 below, report all support for the work reported in this manuscript without time limit. For all other items, the time frame for disclosure is the past 36 months.**

| | | **Name all entities with whom you have this relationship or indicate none (add rows as needed)** | **Specifications/Comments (e.g., if payments were made to you or to your institution)** |
|---|---|---|---|
| | | **Time frame: Since the initial planning of the work** | |
| 1 | All support for the present manuscript (e.g., funding, provision of study materials, medical writing, article processing charges, etc.) **No time limit for this item.** | AB-BIOTICS SA | Payment made to Innovacion y Desarrollo Estrategias en Salud SA de CV. My own company |
| | | **Time frame: past 36 months** | |
| 2 | Grants or contracts from any entity (if not indicated in item #1 above). | None | |
| 3 | Royalties or licenses | None | |

| | | | |
|---|---|---|---|
| 4 | Consulting fees | BioGaia | Consulting fees since 2015 |
| 5 | Payment or honoraria for lectures, presentations, speakers bureaus, manuscript writing or educational events | BioGaia | Honoraria made to me since 2010 acting as speaker |
| 6 | Payment for expert testimony | None | |
| 7 | Support for attending meetings and/or travel | None | |
| 8 | Patents planned, issued or pending | None | |
| 9 | Participation on a Data Safety Monitoring Board or Advisory Board | None | |
| 10 | Leadership or fiduciary role in other board, society, committee or advocacy group, paid or unpaid | None | |
| 11 | Stock or stock options | None | |
| 12 | Receipt of equipment, materials, drugs, medical writing, gifts or other services | None | |
| 13 | Other financial or non-financial interests | None | |

**Please place an "X" next to the following statement to indicate your agreement:**

**X I certify that I have answered every question and have not altered the wording of any of the questions on this form.**

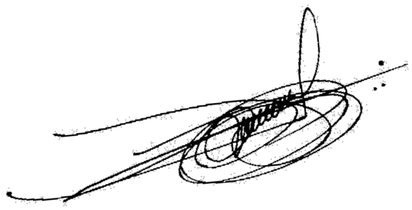

#### ICMJE DISCLOSURE FORM

**Date: May, 10th, 2021**
**Your Name: Tania Gandara-Marti**
**Manuscript Title: Efficacy and safety of probiotic formulation in adult COVID19 outpatients: a randomized, placebo-controlled clinical trial**
**Manuscript number (if known): N.A.**

**In the interest of transparency, we ask you to disclose all relationships/activities/interests listed below that are related to the content of your manuscript. "Related" means any relation with for-profit or not-for-profit third parties whose interests may be affected by the content of the manuscript. Disclosure represents a commitment to transparency and does not necessarily indicate a bias. If you are in doubt about whether to list a relationship/activity/interest, it is preferable that you do so.**

**The following questions apply to the author's relationships/activities/interests as they relate to the current manuscript only.**

**The author's relationships/activities/interests should be defined broadly. For example, if your manuscript pertains to the epidemiology of hypertension, you should declare all relationships with manufacturers of antihypertensive medication, even if that medication is not mentioned in the manuscript.**

**In item #1 below, report all support for the work reported in this manuscript without time limit. For all other items, the time frame for disclosure is the past 36 months.**

| | | Name all entities with whom you have this relationship or indicate none (add rows as needed) | Specifications/Comments (e.g., if payments were made to you or to your institution) |
|---|---|---|---|
| | | **Time frame: Since the initial planning of the work** | |
| 1 | All support for the present manuscript (e.g., funding, provision of study materials, medical writing, article processing charges, etc.) **No time limit for this item.** | AB-BIOTICS SA | Innovación y Desarrollo de Estrategias en Salud SAdeCV which leader the Project received the financial support from AB-BIOTCIS that help to cover the activities related to the develop of the different activities where I participated |
| | | **Time frame: past 36 months** | |
| 2 | Grants or contracts from any entity (if not indicated in item #1 above). | None | |
| 3 | Royalties or licenses | None | |
| 4 | Consulting fees | None | |

| | | |
|---|---|---|
| 5 | Payment or honoraria for lectures, presentations, speakers bureaus, manuscript writing or educational events | None |
| 6 | Payment for expert testimony | None |
| 7 | Support for attending meetings and/or travel | None |
| 8 | Patents planned, issued or pending | None |
| 9 | Participation on a Data Safety Monitoring Board or Advisory Board | None |
| 10 | Leadership or fiduciary role in other board, society, committee or advocacy group, paid or unpaid | None |
| 11 | Stock or stock options | None |
| 12 | Receipt of equipment, materials, drugs, medical writing, gifts or other services | None |
| 13 | Other financial or non-financial interests | None |

**Please place an "X" next to the following statement to indicate your agreement:**

**X I certify that I have answered every question and have not altered the wording of any of the questions on this form.**

**Dra. Tania Gandara-Marti**
